# Seroprevalence of SARS-CoV-2 antibodies and associated factors in health care workers: a systematic review and meta-analysis

**DOI:** 10.1101/2020.10.23.20218289

**Authors:** Petros Galanis, Irene Vraka, Despoina Fragkou, Angeliki Bilali, Daphne Kaitelidou

**Author notes:** Corresponding author: Petros Galanis, PhD, Faculty of Nursing, Center for Health Services Management and Evaluation, National and Kapodistrian University of Athens, 123 Papadiamantopoulou street, GR-11527, Athens, Greece. **Author contributions** P.G, I.V. and D.K. were responsible for the conception and design of the study. P.G, I.V., D.F., A.B. were responsible for the acquisition, analysis and interpretation of data. All the authors drafted the article or revised it critically for important intellectual content, and provided final approval of the version to be submitted. **Funding:** None.

## Abstract

**Background:** Health care workers (HCWs) represent a high risk population for the severe acute respiratory syndrome coronavirus 2 (SARS-CoV-2) infection.

**Aim:** To determine the seroprevalence of SARS-CoV-2 antibodies among HCWs, and to find out the factors that are associated with this seroprevalence.

**Methods:** The Preferred Reporting Items for Systematic Reviews and Meta-Analysis guidelines were applied for this systematic review and meta-analysis. Databases including PubMed/MEDLINE and pre-print services (medRχiv and bioRχiv) were searched from inception up to August 24, 2020.

**Findings:** Forty-nine studies, including 127,480 HCWs met the inclusion criteria. The estimated overall seroprevalence of SARS-CoV-2 antibodies among HCWs was 8.7% (95% CI: 6.7-10.9%). Seroprevalence was higher in studies that were conducted in North America (12.7%) compared to those in Europe (8.5%), Africa (8.2), and Asia (4%). Meta-regression showed that increased sensitivity of antibodies test was associated with increased seroprevalence. The following factors were associated with seropositivity: male gender, Black, Asian, and Hispanic HCWs, work in a coronavirus disease 2019 (COVID-19) unit, patient-related work, frontline health care workers, health care assistants, personal protective equipment shortage, self-reported belief for previous SARS-CoV-2 infection, previous positive polymerase chain reaction test, and household contact with suspected or confirmed COVID-19 patients.

**Conclusion:** The seroprevalence of SARS-CoV-2 antibodies among HCWs is high. Excellent adherence to infection prevention and control measures, sufficient and adequate personal protective equipment, and early recognition, identification and isolation of HCWs that are infected with SARS-CoV-2 are imperative to decrease the risk of SARS-CoV-2 infection.

## Introduction

The severe acute respiratory syndrome coronavirus 2 (SARS-CoV-2) and coronavirus disease 2019 (COVID-19) emerged from the Wuhan, Hubei province, China during December 2019 and the World Health Organization (WHO) declared a world pandemic on 11^th^ March 2020 [1]. As of October 2, 2020, the WHO reported 34,079,542 cases globally and 1,015,963 deaths due to COVID-19 [2].

Health care workers (HCWs) are a high risk group for infection and a recent meta-analysis with 11 studies found that the proportion of HCWs who were SARS-CoV-2 positive among all COVID-19 patients was 10.1% but the severity and mortality among HCWs were lower than COVID-19 patients [3]. This proportion varied substantially among countries i.e. China; 4.2%, Italy; 9% and USA; 17.8% [3]. Lower proportion in China is probably due to implementation of immediate and strong public health interventions e.g. lockdown measures, home isolation, quarantine measures, wearing masks and social (physical) distancing [4].

SARS-CoV-2 and COVID-19 present significant diagnostic issues i.e. serology tests aim to identify previous SARS-CoV-2 infection detecting the presence of SARS-CoV-2 antibodies. Until now, it is known that SARS-CoV-2 antibodies tests are accurate to detect previous SARS-CoV-2 infection if used >14 days after the onset of symptoms and they have very low sensitivity in the first week since symptoms onset [5]. Also, rapid diagnostic tests for SARS-CoV-2 antibodies show a low pooled sensitivity (64.8) and a high pooled specificity (98%) but this meta-analysis suffers from low power and other significant limitations [6].

Knowing seroprevalence of SARS-CoV-2 antibodies among HCWs is important to understand COVID-19 spread among health care facilities and to assess the success of public health interventions. To our knowledge, the overall seroprevalence of SARS-CoV-2 antibodies among HCWs and the associated factors are unknown. Thus, the primary objective of this systematic review and meta-analysis was to determine the seroprevalence of SARS-CoV-2 antibodies among HCWs, while the secondary objective was to find out the factors that are associated with this seroprevalence.

## Methods

### Data sources and strategy

The Preferred Reporting Items for Systematic Reviews and Meta-Analysis (PRISMA) guidelines were applied for this systematic review and meta-analysis [7]. PRISMA checklist is presented in Web Table 1. We searched PubMed/MEDLINE and pre-print services (medRχiv and bioRχiv) from inception up to August 24, 2020. Also, we examined reference lists of all relevant articles and we removed duplicates. We used the following search strategy: (“sars-cov-2 antibodies” OR “COVID-19 antibodies” OR “sars-cov-2” OR “COVID-19” OR antibodies) AND (“health care personnel” OR “healthcare personnel” OR “health-care personnel” OR “health care workers” OR “health-care workers” OR “healthcare workers” OR “healthcare staff” OR “health care staff” OR “health-care staff” OR “medical staff”).

**Table 1.**
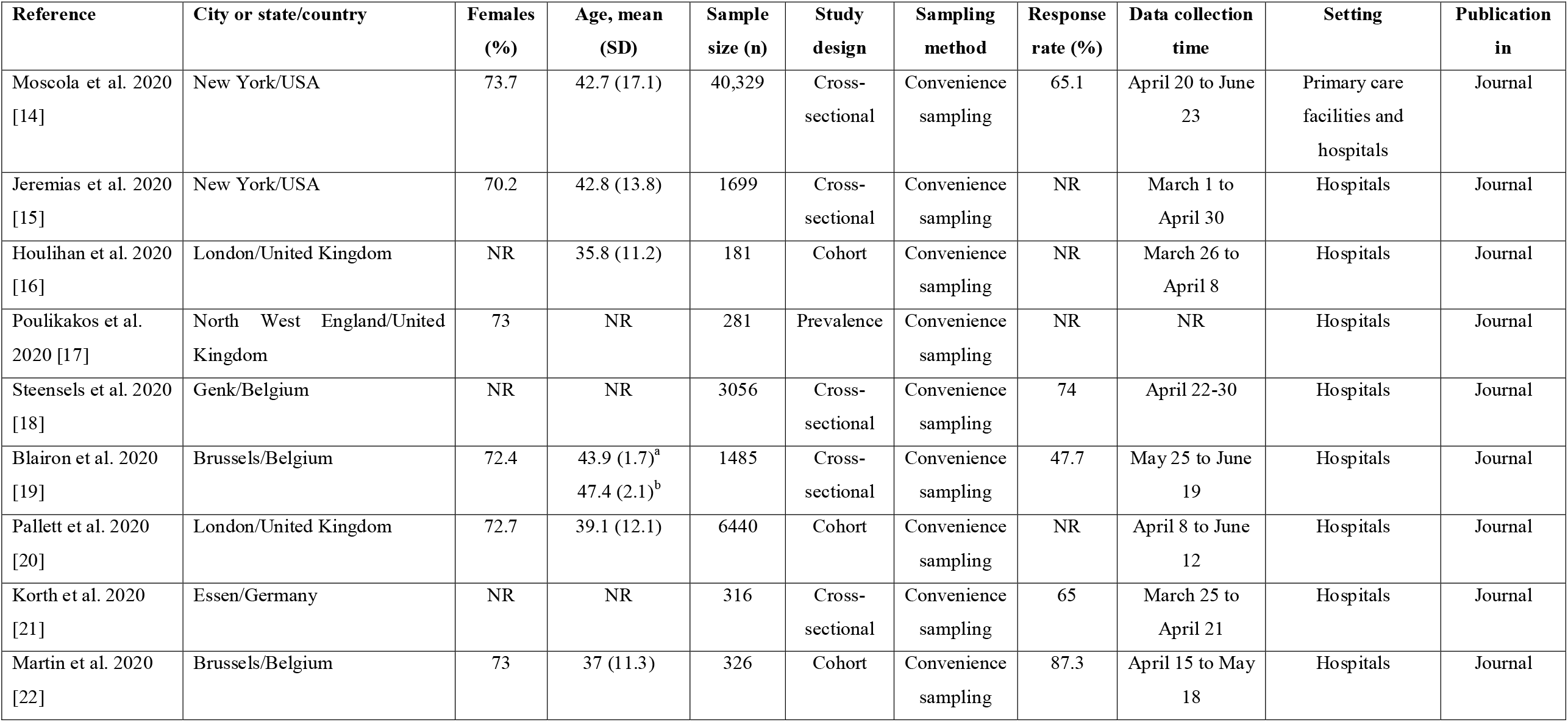
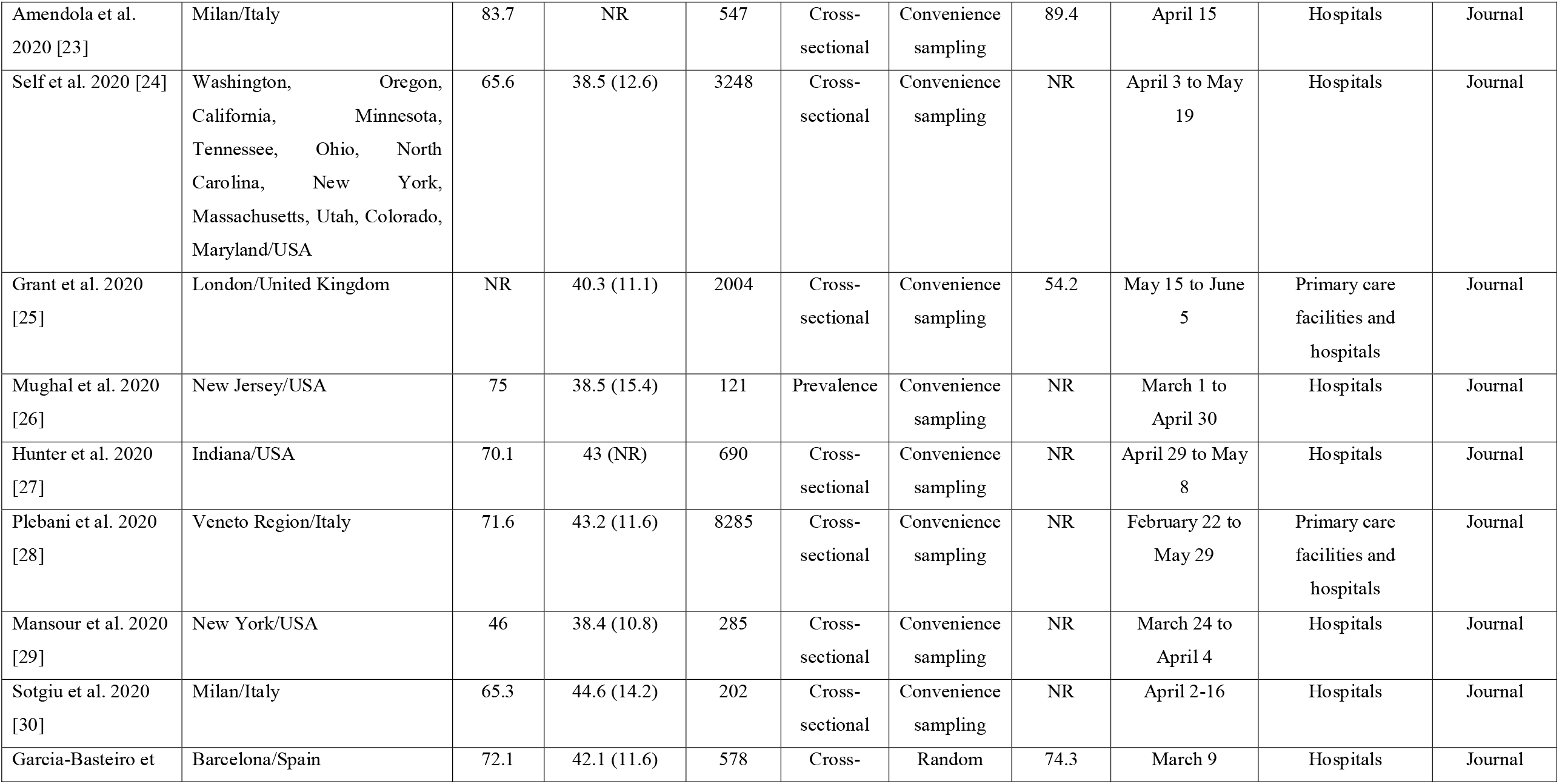
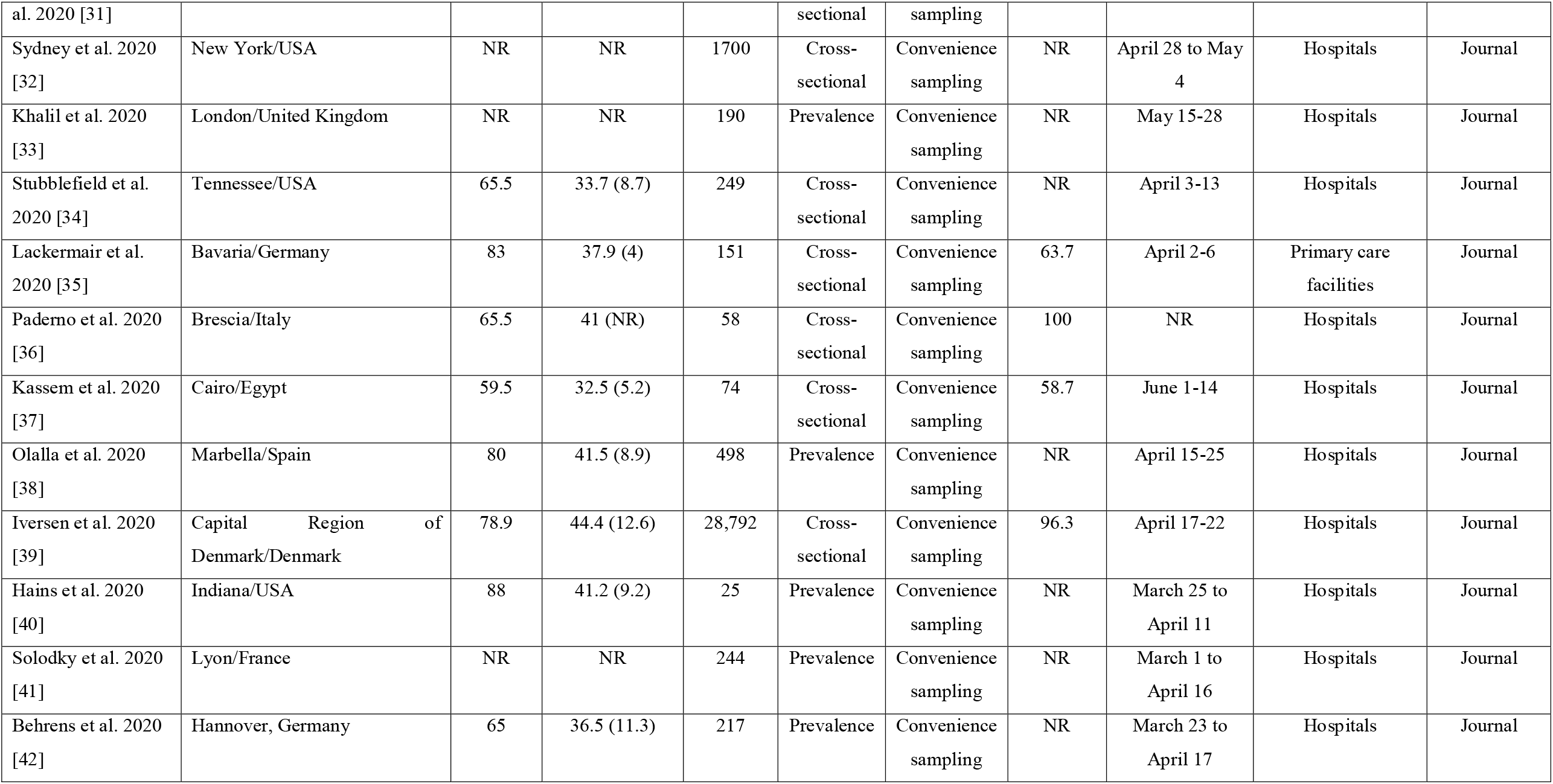
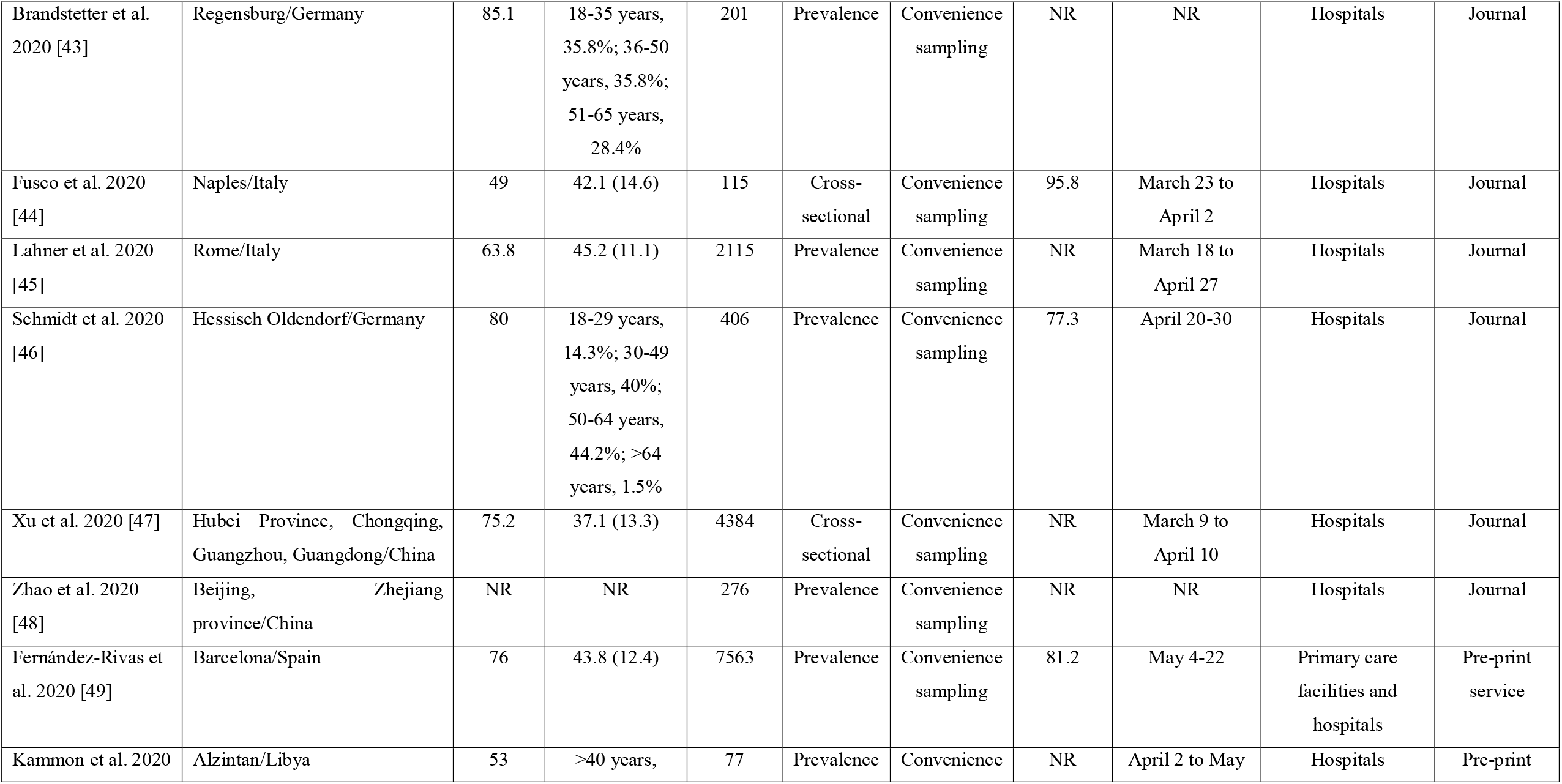
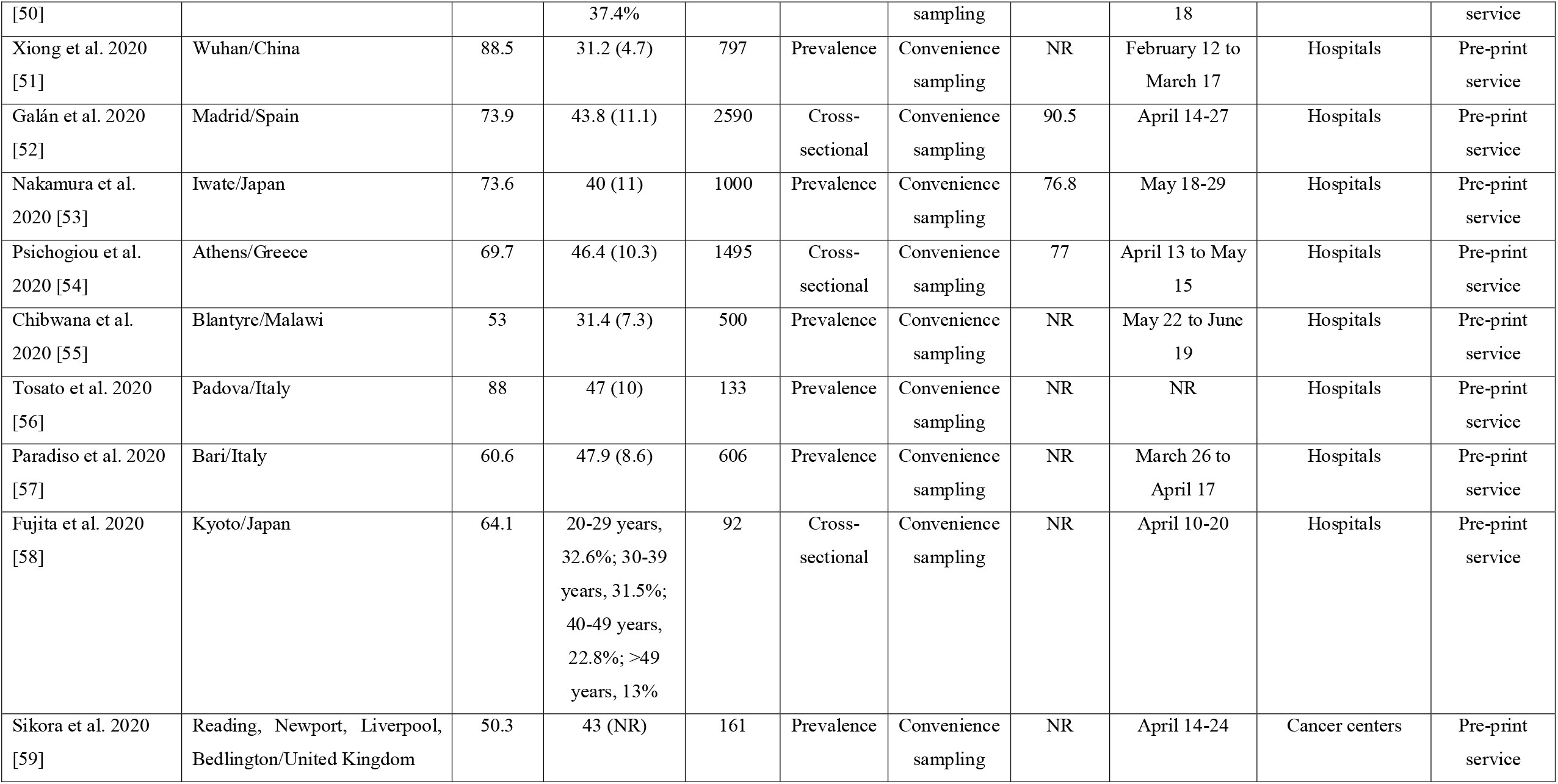
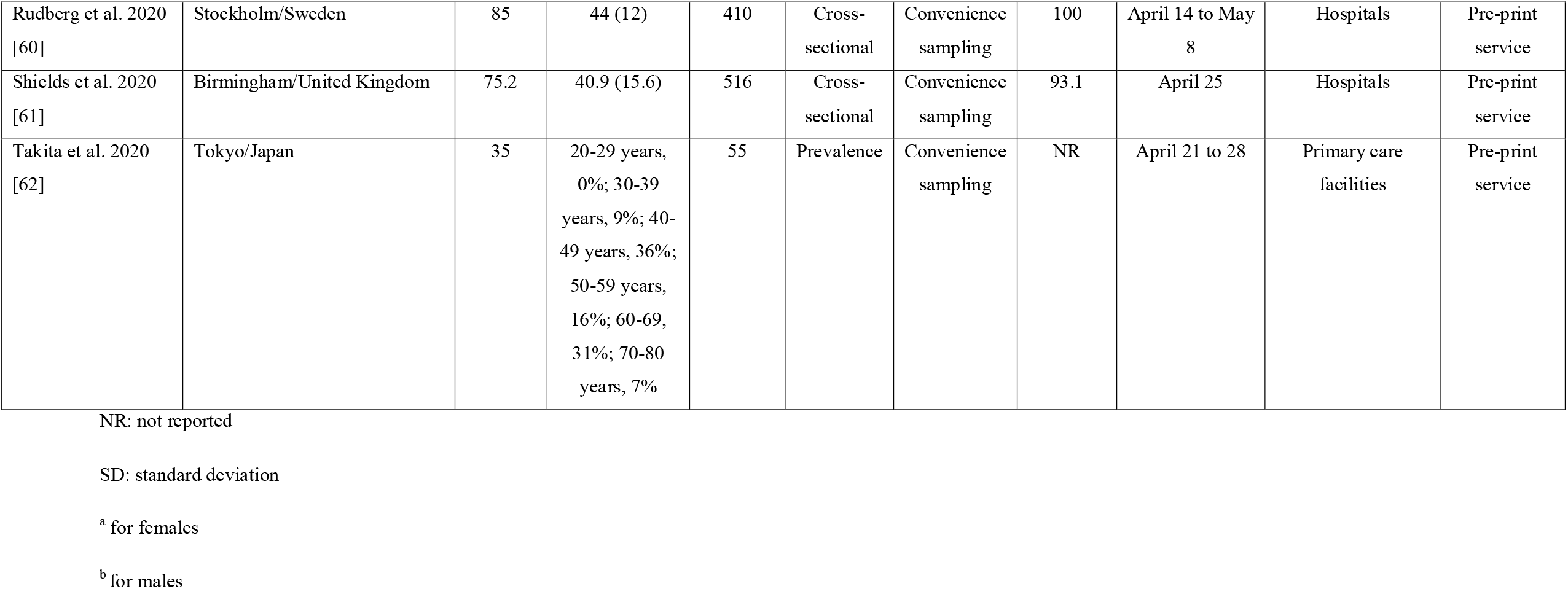
Main characteristics of the studies included in the systematic review and meta-analysis.

### Selection and eligibility criteria

Two independent review authors performed study selection and a third, senior author resolved the discrepancies. We included all studies that were written in English, except case reports. Also, we included studies that reported the seroprevalence of SARS-CoV-2 antibodies among HCWs and the associated factors. We searched for any serology test (e.g. ELISA, CLIA, etc.) detects SARS-CoV-2 antibodies (IgA, IgG, and IgM) in all HCWs. Moreover, we included studies that were performed under screening settings and HCWs were neither selected for participation based on previous exposure to SARS-CoV-2 nor symptoms.

### Data extraction and quality assessment

Data collected included study characteristics such as authors, location, data collection time, sample size, setting, study design, antibodies test, sensitivity and specificity for the antibodies test, number of HCWs with SARS-CoV-2 antibodies, factors associated with the seroprevalence of SARS-CoV-2 antibodies, and the level of analysis (univariate or multivariable).

The quality of the studies was assessed with the Joanna Briggs Institute critical appraisal tools where a 9-point scale is used for prevalence studies, an 8-point scale for cross-sectional studies and an 11-point scale for cohort studies [8]. In prevalence studies, a score of 8-9 points indicates good quality, a score of 5-7 points indicates moderate quality and a score ≤4 indicates poor quality. In cross-sectional studies, a score of 7-8 points indicates good quality, a score of 4-6 points indicates moderate quality and a score ≤3 indicates poor quality. In cohort studies, a score of 9-11 points indicates good quality, a score of 5-8 points indicates moderate quality and a score ≤4 indicates poor quality.

### Statistical analysis

For each study we extracted the total number of HCWs and the number of HCWs that were positive for SARS-CoV-2 antibodies. The seroprevalence and the 95% confidence interval (CI) were calculated for each included study. We transformed seroprevalences with the Freeman-Tukey Double Arcsine method before pooling [9]. Between-studies heterogeneity was assessed by the Hedges Q statistic and I^2^ statistics. Statistical significance for the Hedges Q statistic is set at p-value < 0.1, while I^2^ values higher than 75% indicates high heterogeneity [10]. We applied a random effect model to estimate pooled seroprevalence since the heterogeneity between results was very high [10–11]. We considered studies quality, sample size, sensitivity and specificity for the antibodies tests, publication type (journal or pre-print service) and the continent that studies were conducted as pre-specified sources of heterogeneity and we explored them with subgroup analysis and meta-regression analysis. Also, we performed a leave-one-out sensitivity analysis by removing one study at a time to determine the influence of each study on the overall prevalence. We used a funnel plot and the Egger’s test to assess the publication bias. P-value < 0.05 for the Egger’s test indicates publication bias [12]. We did not perform meta-analysis for the factors that are associated with the seroprevalence of SARS-CoV-2 antibodies since the data were very scarce. Statistical analysis was performed with OpenMeta[Analyst] [13].

## Results

### Identification and selection of studies

Flowchart of the literature search is summarized in PRISMA format (Figure 1). Initially, we identified 3632 potential records through PubMed and 103 records through preprint servers for health sciences i.e. medRχiv and bioRχiv removing duplicates. After the screening of the titles and abstracts, we removed 3684 records and we added 12 more records found by the reference lists scanning. Finally, we included 49 studies in this meta-analysis that met our inclusion criteria.

**Figure 1.**
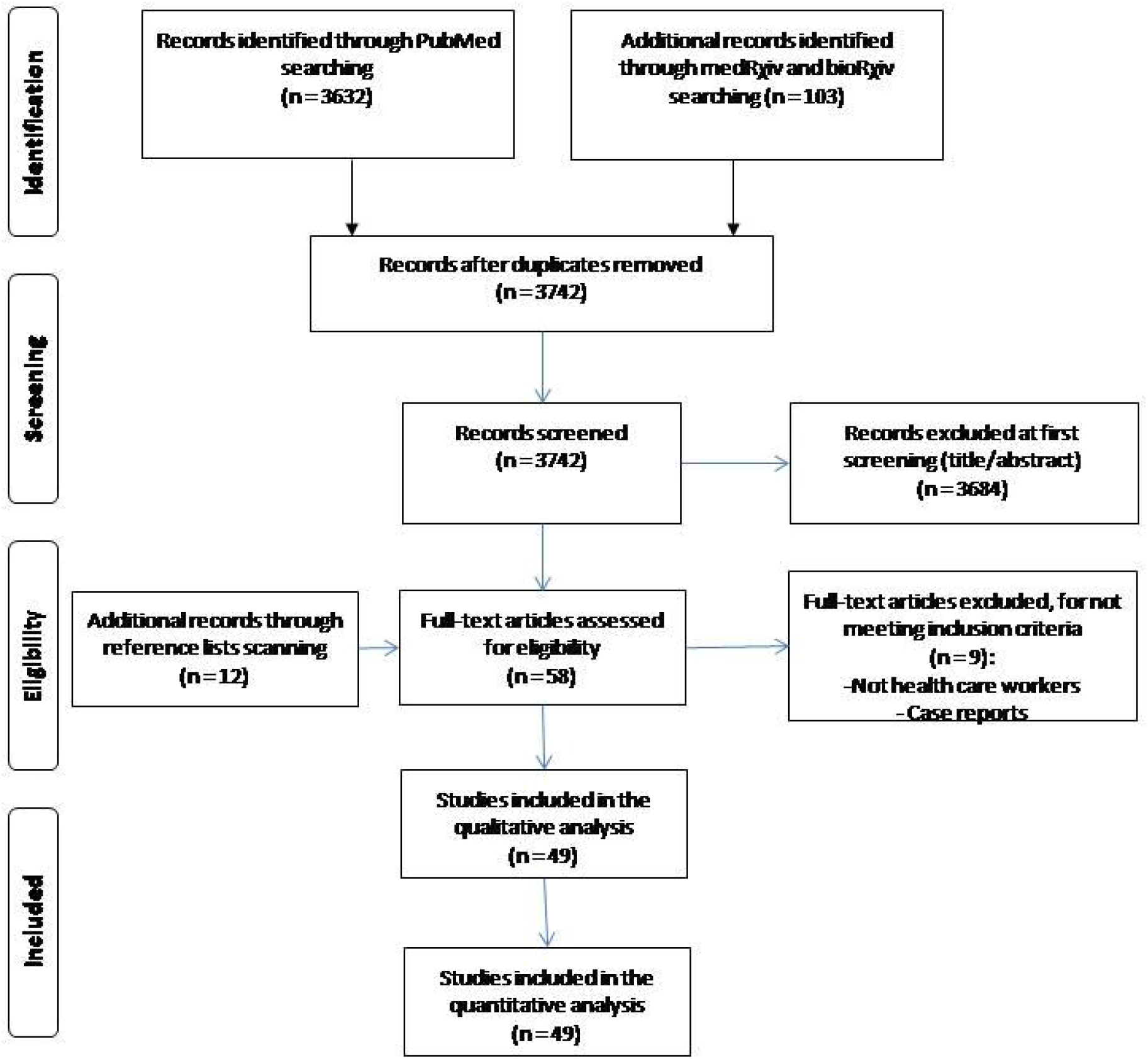
Flowchart of the literature search according to the Preferred Reporting Items for Systematic Reviews and Meta-Analysis.

### Characteristics of the studies

Main characteristics of the 49 studies included in our systematic review and meta-analysis are shown in Table 1. A total of 127,480 HCWs were included in this systematic review and meta-analysis. Forty-nine studies [14–62] reported data regarding the seroprevalence of SARS-CoV-2 antibodies among HCWs and 27 studies [14, 15, 18, 19, 21–25, 27–32, 34–37, 39, 44, 47, 52, 54, 58, 60, 61] investigated factors for SARS-CoV-2 antibodies positivity.

The majority of studies was conducted in Europe (n=31), and then in North America (n=9), Asia (n=6), and Africa (n=3). In particular, nine studies was conducted in USA [14, 15, 24, 26, 27, 29, 32, 34, 40], eight studies in Italy [23, 28, 30, 36, 44, 45, 56, 57], seven studies in United Kingdom [16, 17, 20, 25, 33, 59, 61], five studies in Germany [21, 35, 42, 43, 46], four studies in Spain [31, 38, 49, 52], and three studies in and Japan [53, 58, 62], Belgium [18, 19, 22] and China [47, 48, 51]. Twenty-nine studies did not report the response rate [15–17, 20, 24, 26–30, 32–34, 38, 40–43, 45, 47, 48, 50, 51, 55–59, 62], nine studies did not report HCWs’ age [17, 18, 21, 23, 32, 33, 36, 41, 48], eight studies did not report sex distribution [16, 18, 21, 25, 32, 33, 41, 48] and five studies did not report data collection time [17, 36, 43, 48, 56]. Females’ percentage ranged from 35% [62] to 88.5% [51] and was higher in 41 studies, while in three studies males’ percentage was higher. Mean age of HCWs ranged from 31.2 [51] years to 47.9 years [57], while sample size ranged from 25 [40] to 40,329 HCWs [14]. Regarding the study design, 26 studies were cross-sectional studies [14, 15, 18, 19, 21, 23–25, 27–32, 34–37, 39, 44, 47, 52, 54, 58, 60, 61], 20 studies were prevalence studies [17, 26, 33, 38, 40–43, 45, 46, 48–51, 53, 55, 56, 57, 59, 62], and three studies were cohort studies [16, 20, 22]. All studies except one [31] used a convenience sample, while response rate ranged from 47.7% [19] to 100% [36, 60]. Forty-two studies were conducted in hospitals [15–24, 26, 27, 29–48, 50–62], four studies in primary care facilities and hospitals [14, 25, 28, 49], two studies in primary care facilities [35, 62] and one study in cancer centers [59]. Thirty-five studies were published in journals [14–48], while 14 studies in pre-print services [49–62].

Validity assessment (sensitivity and specificity) for the antibodies tests used in the included studies according to the manufacturers data are presented in Web Table 2. Sensitivity ranged from 50% to 100%, while specificity from 80.5% to 100%.

**Table 2.**
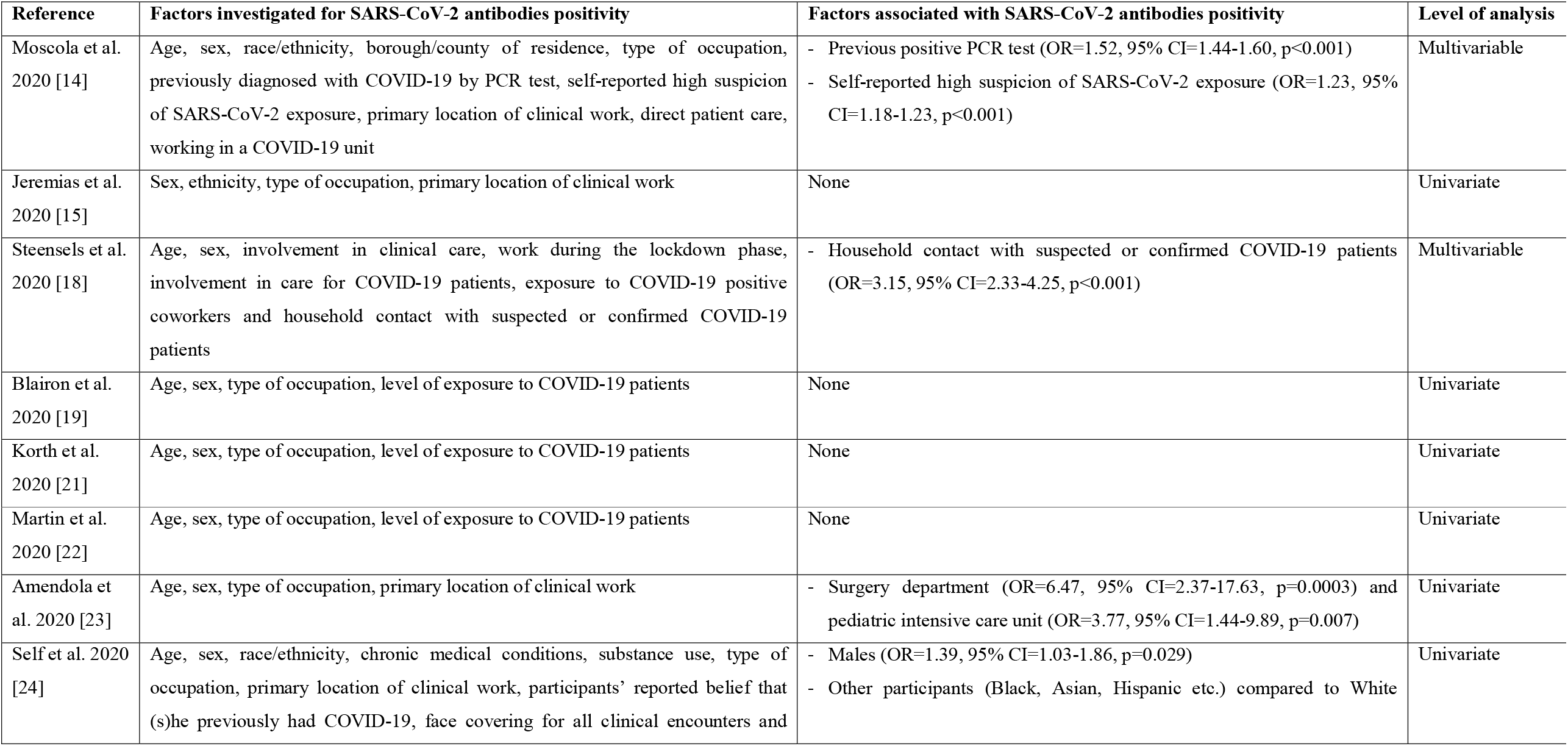
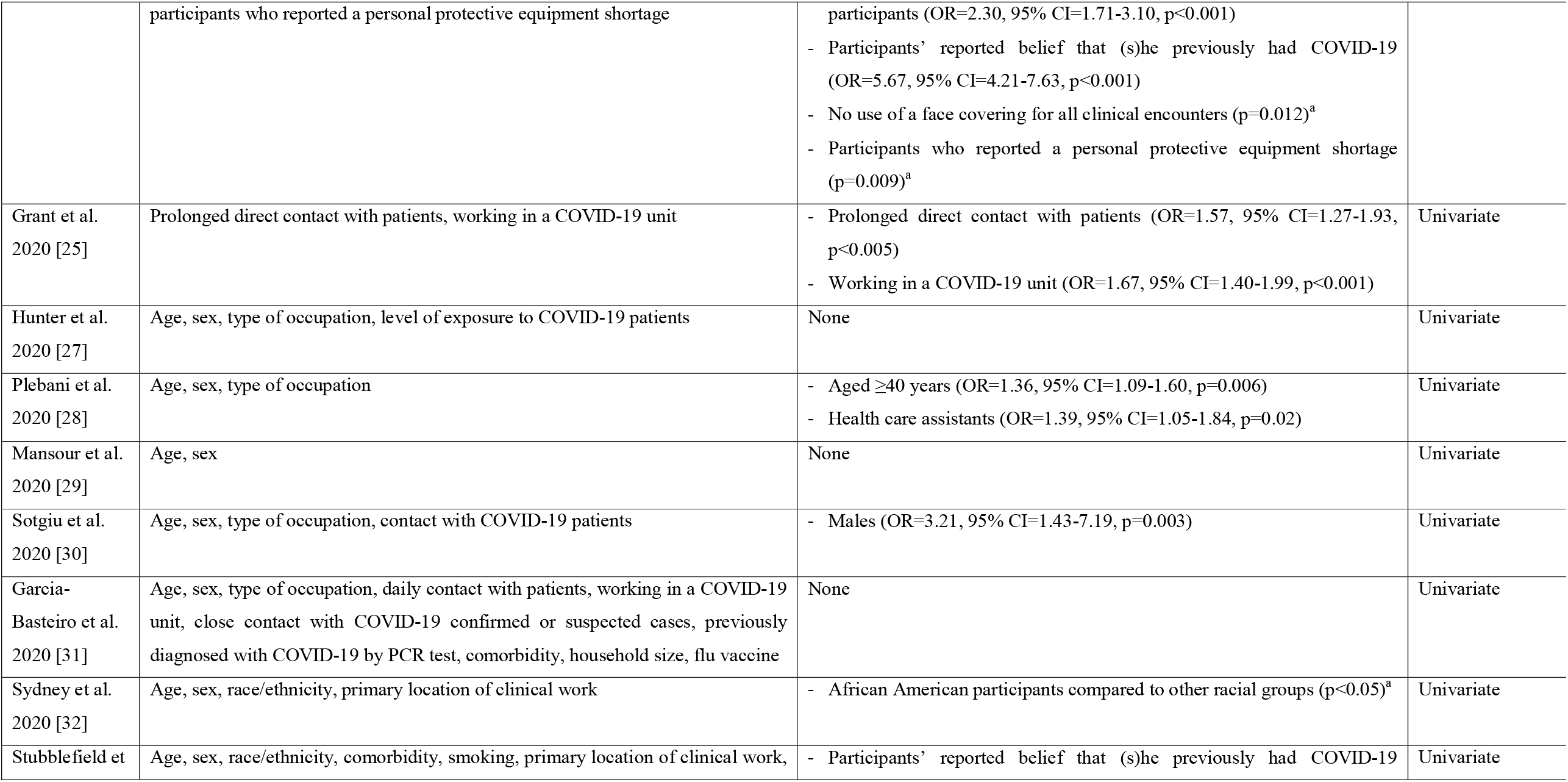
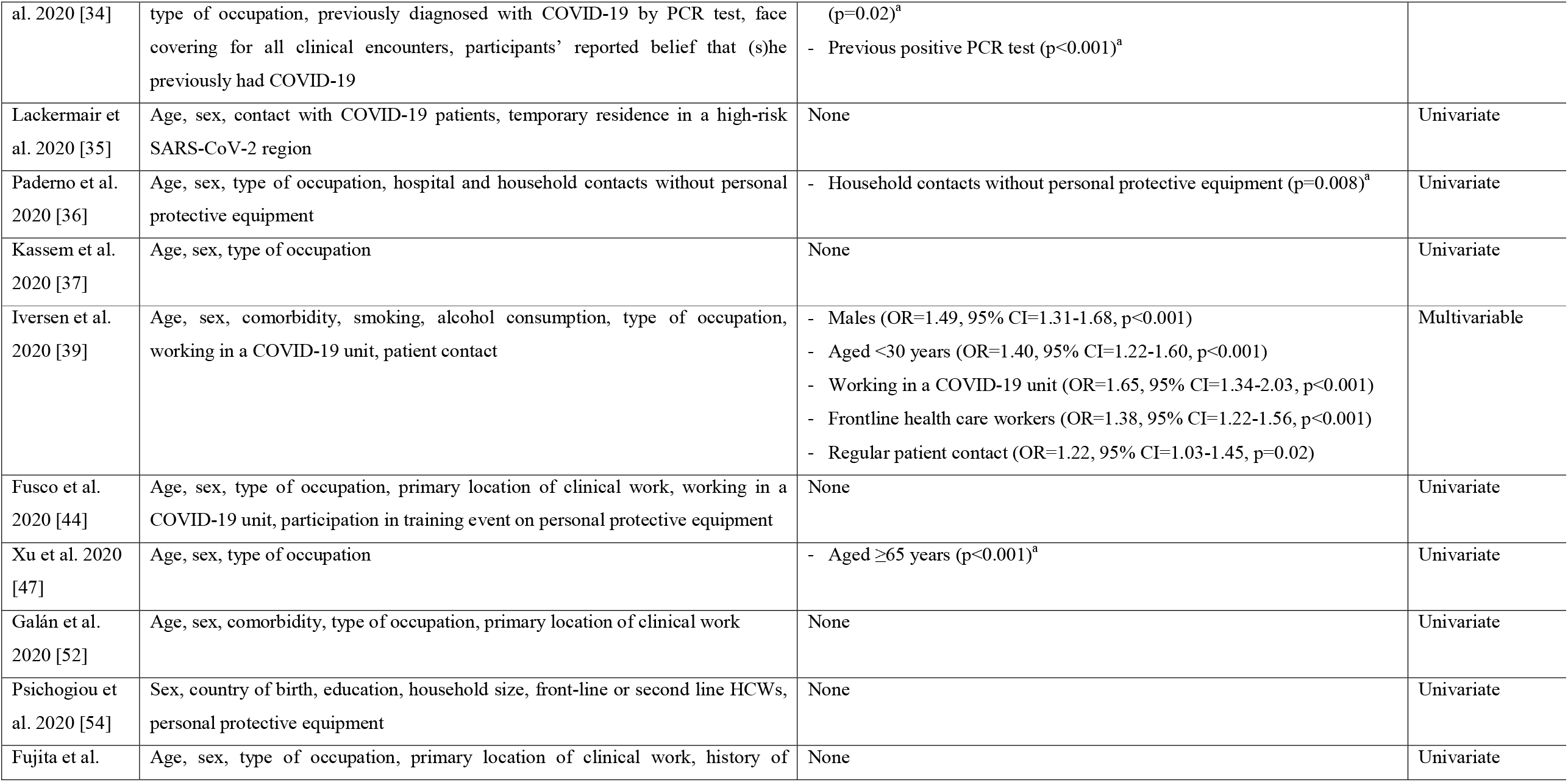
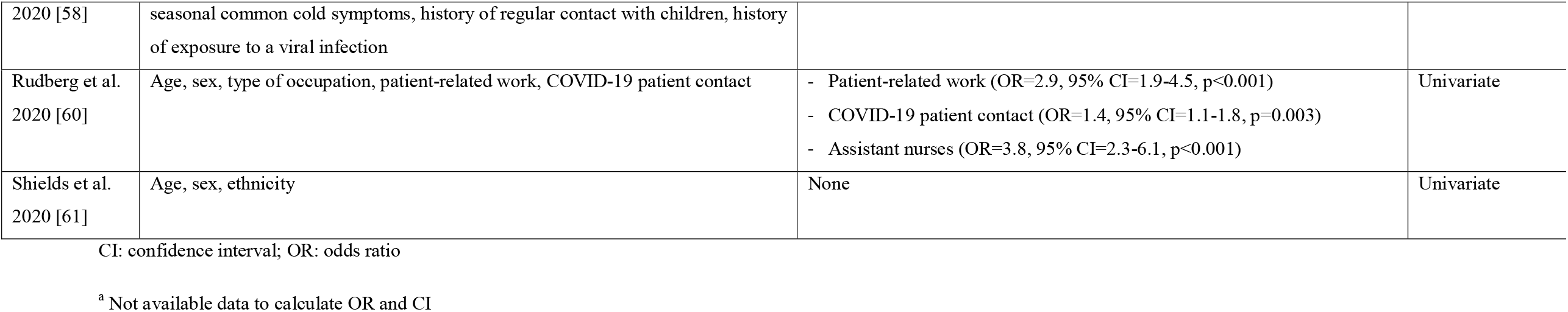
Studies that investigated factors associated with SARS-CoV-2 antibodies positivity among health care workers.

### Quality assessment

Quality assessment of prevalence studies, cross-sectional studies and cohort studies are shown in Web Tables 3, 4, and 5 respectively. Quality was moderate in 37 studies, good in 10 studies, and poor in two studies. Regarding prevalence studies, 16 were at moderate risk of bias, three were at low risk and one was at high risk. Moreover, 20 cross-sectional studies were at moderate risk of bias, five were at low risk and one was at high risk. Two cohort studies were at low risk of bias and one was at moderate risk.

### Meta-analysis of the seroprevalence

We applied a random effect model to estimate pooled prevalence since the heterogeneity between results was very high (I^2^=99.34, p-value for the Hedges Q statistic < 0.001). The estimated overall seroprevalence of SARS-CoV-2 antibodies among HCWs was 8.7% (95% CI: 6.7-10.9%) (Figure 2). Seroprevalence among studies ranged from 0% to 45.3%.

**Figure 2.**
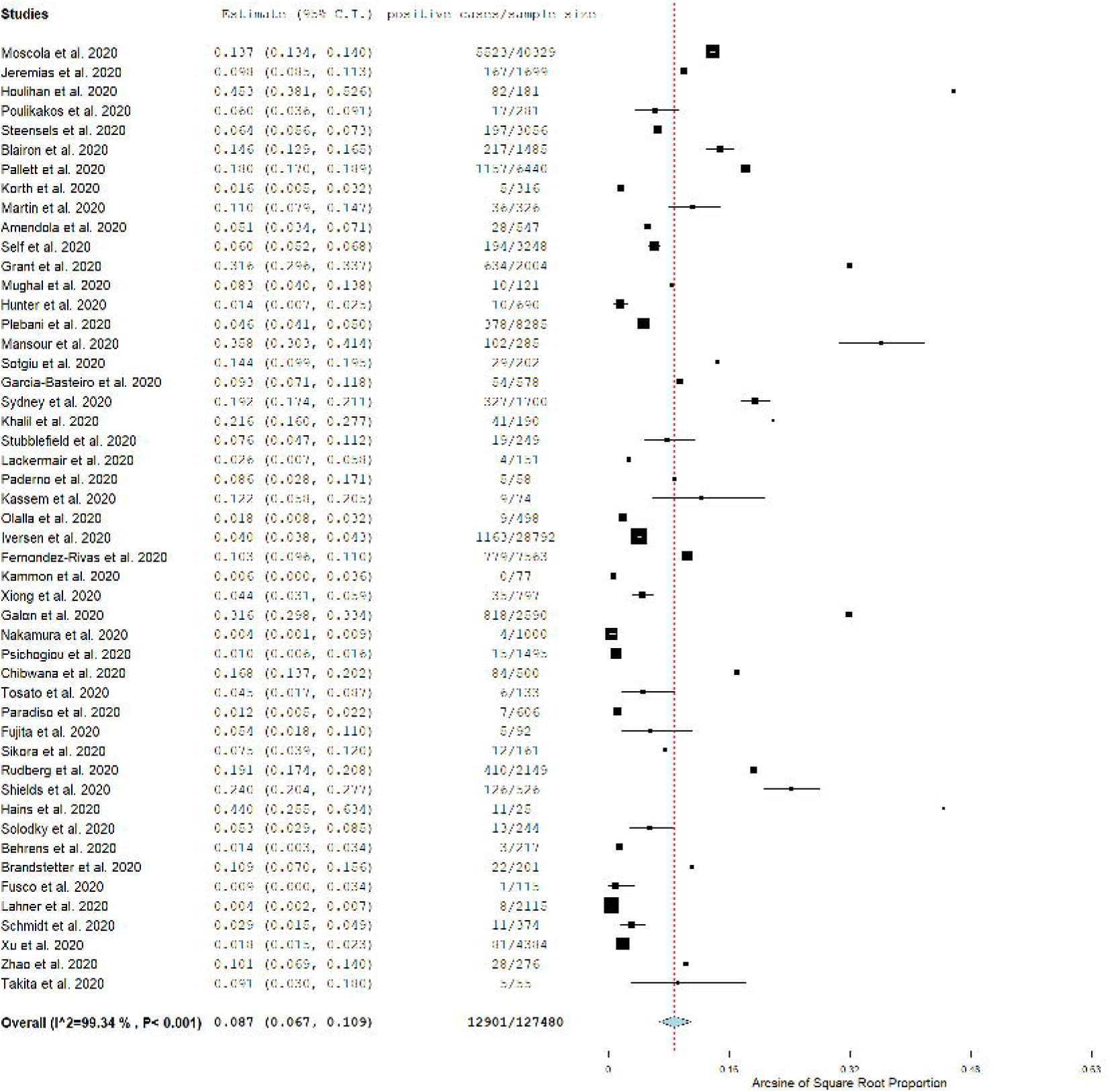
Forest plot of the seroprevalence of SARS-CoV-2 antibodies with corresponding 95% confidence intervals. The size of the black boxes is positively proportional to the weight assigned to studies, and horizontal lines represent the 95% confidence intervals according to random effects analysis.

### Subgroup and meta-regression analysis

According to subgroup analysis, seroprevalence of SARS-CoV-2 antibodies was higher for the studies with poor quality (11.6% [95% CI: 0.7-32.7%]) compared to those with moderate quality (8.8% [95% CI: 6.0-12%]) and good quality (7.9% [95% CI: 4.1-12.8%]). Moreover, seroprevalence was higher for the studies that were published in journals (9% [95% CI: 6.7-11.6%]) compared to those in pre-print services (7.7% [95% CI: 3.4-13.4%]). Seroprevalence was higher in studies that were conducted in North America (12.7% [95% CI: 8.6-17.5%]) compared to those in Europe (8.5% [95% CI: 5.8-11.6%]), Africa (8.2% [95% CI: 0.8-22.3%]), and Asia (4% [95% CI: 1.8-7.1%]). Meta-regression showed that increased sensitivity of antibodies test was associated with increased seroprevalence (coefficient = 0.004 [95% CI: 0.0001-0.009], p=0.038). Moreover, seroprevalence was independent of the sample size (p=0.65) and the specificity (p=0.20).

### Sensitivity analysis

A leave-one-out sensitivity analysis showed that no single study had a disproportional effect on the overall seroprevalence, which varied between 8.2% (95% CI: 6.2-10.3%), with Hoolihan et al. [16] excluded, and 9.0% (95% CI: 6.9-11.2%), with Nakamura et al. [53] excluded (Web Figure 1).

### Publication bias

Egger’s test (p=0.0001) and the asymmetrical shape of the funnel plot (Web Figure 2) implied potential publication bias.

### Factors associated with SARS-CoV-2 antibodies positivity

Twenty-seven studies [14, 15, 18, 19, 21–25, 27–32, 34–37, 39, 44, 47, 52, 54, 58, 60, 61] investigated factors associated with SARS-CoV-2 antibodies positivity and 13 studies found associations [14, 18, 23–25, 28, 30, 32, 34, 36, 39, 47, 60] (Table 2). Twenty-four studies [15, 19, 21–25, 27–32, 34–37, 44, 47, 52, 54, 58, 60, 61] used univariate analysis, while three studies [14, 18, 39] used multivariable regression analysis.

Three studies found [24, 30, 39] that males have more frequently detectable SARS-CoV-2 antibodies with odds ratios (OR) ranging from 1.39 to 3.21. Results regarding age were controversial since SARS-CoV-2 antibodies positivity was associated with HWCs aged <30 years (OR=1.40, 95% CI=1.22-1.60) [39], HWCs aged ≥40 years (OR=1.36, 95% CI=1.09-1.60) [28], and HWCs aged ≥65 years (p<0.001) [47]. A significantly higher percentage of SARS-CoV-2 antibodies found among African American HCWs (p<0.05) [32] and Black, Asian, and Hispanic HCWs compared to White participants (OR=2.30, 95% CI=1.71-3.10, p<0.001) [24].

Three studies [25, 39, 60] found a significantly higher probability of a positive SARS-CoV-2 antibodies test in HCWs working in a COVID-19 unit with ORs ranging from 1.4 to 1.67. Similar results found for HCWs with patient-related work (ORs range from 1.22 to 2.9) [25, 39, 60] and frontline health care workers (OR=1.38, 95% CI=1.22-1.56) [39]. Moreover, Self et al. [24] found that HCWs in surgery department (OR=6.47, 95% CI=2.37-17.63) and pediatric intensive care unit (OR=3.77, 95% CI=1.44-9.89, p=0.007) had a significantly higher percentage of SARS-CoV-2 antibodies. Two studies [28, 60] found that SARS-CoV-2 antibodies positivity was higher among health care assistants (OR=1.39, 95% CI=1.05-1.84 and OR=3.8, 95% CI=2.3-6.1). Self et al. [24] found that no use of a face covering for all clinical encounters (p=0.012) and personal protective equipment shortage (p=0.009) increase the probability of a positive SARS-CoV-2 antibodies test in HCWs.

Three studies [14, 24, 34] found that HCWs self-reported belief that (s)he previously had COVID-19 (ORs range from 1.23 to 5.67) is associated with SARS-CoV-2 antibodies positivity. Similar results found for HCWs with a previous positive Polymerase Chain Reaction (PCR) test (OR=1.52, 95% CI=1.44-1.60 in one study [14] and p<0.001 in another study [34]). Also, two studies [18, 36] found that household contact with suspected or confirmed COVID-19 patients increases the probability of a positive SARS-CoV-2 antibodies test in HCWs (OR=3.15, 95% CI=2.33-4.25 in one study [18] and p=0.008 in another study [36]).

## Discussion

To our knowledge, this is the first systematic review and meta-analysis that estimates the overall seroprevalence of SARS-CoV-2 antibodies among HCWs in screening settings. We found that the overall seroprevalence was 8.7% with a wide range among studies from 0% to 45.3%. Population-based and community-based studies in USA showed a great variability in seroprevalence of SARS-CoV-2 antibodies from 1.1% to 14.4% [63–67]. Similar studies in Europe [68–70] and China [71] found very different seroprevalence in general population ranging from 0.23% to 10.9%. These differences in seroprevalence among studies may be attributable to several reasons, e.g. different study populations, different antibodies tests with variation in sensitivity and specificity, different study designs, different lockdown and quarantine measures, different data collection time period etc. Moreover, according to our subgroup analysis, seroprevalence of SARS-CoV-2 antibodies was higher for the studies with poor quality (11.6%) compared to those with moderate quality (8.8%) and good quality (7.9%) indicating that difference in studies quality could be also a significant reason for difference in seroprevalence.

Our subgroup analysis identified that seroprevalence was higher in studies that were conducted in North America (12.7%) compared to those in Europe (8.5%), Africa (8.2%), and Asia (4%). This finding is in accordance with a meta-analysis [3] that found that the overall proportion of HCWs who are SARS-CoV-2 positive among all COVID-19 patients is lower in China (4.2%) than in USA (17.8%) and Europe (9%). This might be explained due to the good adherence to infection prevention and control measures and the appropriate use of personal protective equipment among HCWs in China. Also, USA and Europe seem to be unprepared to handle the surge of patients that led to the severe shortage in the personal protective equipment, while USA and most of the countries in Europe (with significant exceptions such as Germany and Greece) took action too late [72]. For example, according to reports in United Kingdom and Italy, HCWs experienced extreme situations during COVID-19 pandemic wearing paper face masks and plastic aprons instead of appropriate masks, visors, and gowns [73, 74]. In our meta-analysis, seroprevalence in studies in United Kingdom (n=7) and Italy (n=8) was higher (10.3%) than the overall seroprevalence (8.4%), while seroprevalence in studies in Germany (n=5) and Greece (n=2) was quite lower (2.2%) than the overall seroprevalence. On the other hand, China has already controlled rapidly and efficiently the Severe Acute Respiratory Syndrome (SARS) epidemic that broke out in 2003 [75, 76]. Thus, China immediately adopts the lessons from handling the SARS epidemic in the case of COVID-19 pandemic applying effective measures, e.g. early case identification and isolation, active large-scale surveillance of individuals even with smartphone application, tracing and quarantining of COVID-19 contacts, temperature screening in public places, physical distancing, travelers screening, street camera system for identification of individuals without a mask or showing symptoms etc. [71, 77, 78]. Moreover, some hospitals in China implemented a tactical training protocol for all aspects of COVID-19 that result in very low infection rate among HCWs even among frontline HCWs in Wuhan [79].

We found that seropositivity was higher for HCWs with patient-related work [25, 39, 60], and frontline health care workers [39]. Grant et al. [25] and Rudberg et al. [60] found that seropositivity of HCWs is much higher than this of general population of London and Stockholm respectively, indicating an occupational health risk among HCWs. Several studies emphasize the risk of occupational transmission of SARS-CoV-2 among HCWs since the HCWs are at the frontline response to the COVID-19 and would be more prone to viral transmission [73, 80–84]. Increased HCWs exposure to SARS-CoV-2 may be attributable mainly to patient-to-HCW transmission and HCW-to-HCW transmission due to the personal protective equipment shortage, poor adherence to infection prevention and control measures, and space constraints in hospitals. Additionally, we found that SARS-CoV-2 antibodies positivity was higher among health care assistants [28, 60] a finding that further reinforces a patient-related transmission of SARS-CoV-2 to HCWs since this occupation involves the most patient near contact.

In our systematic review, the seroprevalence was higher among HCWs working in a COVID-19 unit [25, 39, 60]. It is clear that HCWs with COVID-19 patient contact have represented a high-risk group for SARS-CoV-2 infection especially during the first months of COVID-19 pandemic where the knowledge, the control measures and the personal protective equipment were limited. Also, Self et al. [24] found that no use of a face covering for all clinical encounters and personal protective equipment shortage increase the probability of a positive SARS-CoV-2 antibodies test in HCWs. Thus, personal protective equipment supplies for HCWs at hospitals are a necessary tool against COVID-19, while universal masking is of utmost importance since decreases rate of SARS-CoV-2 infection among HCWs [85]. Optimal personal protective equipment is still unknown but rigorous application of personal protective equipment measures and absolute adherence to all infection prevention and control measures are crucial to reduce SARS-CoV-2 nosocomial transmissions [86–89]. Interestingly, Grant et al. [25] found that seropositivity was lower among intensive care unit HCWs. Several reasons could explain this finding such as the enhanced personal protective equipment for intensive care unit HCWs, the fact that the intubated patients are ventilated on a closed circuit, and the fact that COVID-19 patients who require admission on an intensive care unit are often admitted around day 10 of the natural history of their illness [90], by which point viral loads of patients tend to decrease [91].

According to our review, household contact with suspected or confirmed COVID-19 patients is associated with positive SARS-CoV-2 antibodies test in HCWs [18, 36]. Also, HCWs self-reported belief that (s)he previously had COVID-19 is associated with SARS-CoV-2 antibodies positivity [14, 24, 34]. HCWs are exposed to SARS-CoV-2 not only in clinical settings but also in their house or in social meetings, joint meals, and office spaces with friends or colleagues. In fact, as community transmission increases, the risk of SARS-CoV-2 exposure for HCWs is higher outside of the clinical settings through household contacts with infected COVID-19 patients or interaction with others in areas with active, unmitigated transmission [92–94].

We found that a previous positive PCR test increases the probability of a positive SARS-CoV-2 antibodies test in HCWs [14, 34]. SARS-CoV-2 antibodies tests identify previous SARS-CoV-2 infection but many issues are still controversial. For example, the sensitivity of these tests is too low in the first week since symptom onset but increases ≥15 days after the onset of symptoms [5]. Also, the duration of antibody rises is unknown since the data >35 days after the onset of symptoms are very scarce [5]. Moreover, it is currently unknown whether antibody titers correlate with protective immunity against reinfection and if antibody responses differ significantly in asymptomatic individuals and individuals with mild or severe COVID-19 [95, 96]. Variation in the validity of commercial SARS-CoV-2 antibodies tests, cross-reactivity between SARS-CoV-2 and other coronaviruses, and confusion regarding the possible role of SARS-CoV-2 antibodies as biomarkers of protective immunity or past infection increase the uncertainty about the utility of SARS-CoV-2 antibodies tests in clinical practice [5, 97, 98]. In any case, SARS-CoV-2 antibodies tests seem to be an additional tool against COVID-19 and their utility will be expanded as additional data give us a better understanding of the pros and cons of these tests. Also, universal screening for SARS-CoV-2 in high-risk units in hospitals could help to identify asymptomatic HCWs resulting on self-isolation for the appropriate time [22].

We identified that seropositivity was higher among African American HCWs [32] and Black, Asian, and Hispanic HCWs compared to White participants [24]. This finding is confirmed by studies in general populations where a higher percentage of SARS-CoV-2 antibodies found among Blacks [67, 99] and Hispanics [67]. According to a preliminary analysis of Cook et al. [100], until 12 April, 106 HCWs died in the United Kingdom with COVID-19 and 64.2% (n=68) of them were from the Black, Asian and Minority Ethnic community. Moreover, Gould and Wilson [101] found that Black workers experience higher SARS-CoV-2 seroprevalence than Whites. This disparity may be attributable to several reasons, e.g. work conditions, economic inequality, high population density, limited access to health care services, health insurance etc. There is a need for strategies tailored to the culture of minority groups and organized by local minority leaders, who can mobilize individuals to participate in screening tests, and tracing and quarantining of COVID-19 contacts to avoid additional SARS-CoV-2 infections in minority groups [102].

Our study has several limitations. First, 14 out of 49 included studies were published in pre-print services which do not apply peer-review process. Nevertheless, we assessed studies quality and we performed subgroup analysis according to publication type (journal or pre-print service) and studies quality. Second, the heterogeneity between results was very high. We performed random effects model and subgroups analysis to overcome this limitation. Third, seroprevalence reported in studies could be underestimated or overestimated depending on the applied antibody test. Validity (sensitivity and specificity) of the antibodies tests have not been reported in most of the included studies. We performed meta-regression analysis with sensitivity and specificity of the antibodies tests according to the manufacturers’ data as the moderator variables in order to overcome this limitation. Fourth, time between exposure and antibody testing in studies is unknown and seropositivity may have been missed if testing was too early. This systematic bias could result in an underestimation of the seroprevalence. Last, the data regarding the factors that are associated with the seroprevalence of SARS-CoV-2 antibodies were very scarce and we cannot perform meta-analysis; thus a qualitative approach was applied to assess these factors.

## Conclusion

Sseroprevalence of SARS-CoV-2 antibodies among HCWs is high indicating that HCWs represent a population with considerable risk contracting SARS-CoV-2 infection. Absolute adherence to infection prevention and control measures, sufficient and adequate personal protective equipment, and early recognition, identification and isolation of HCWs that are infected with SARS-CoV-2 are imperative to decrease the risk of SARS-CoV-2 infection. Moreover, seroprevalence studies among HCWs could add significant information regarding the level of exposure among HCWs, the identification of high-risk departments in hospitals, the measurement of COVID-19 spread, the success of interventions, and the understanding of asymptomatic transmission of SARS-CoV-2 in clinical settings. Given the limitations both of our review and the studies that were included, and that the COVID-19 pandemic is still evolving, there is a need for further high-quality studies.

**Web Table 1**. PRISMA Checklist.

**Web Table 2**. Validity assessment (sensitivity and specificity) for the antibodies tests used in the included studies according to the manufacturers data.

**Web Table 3**. Quality of prevalence studies.

**Web Table 4**. Quality of cross-sectional studies.

**Web Table 5**. Quality of cohort studies.

## Supporting information

Web Tables

## Data Availability

All data could be available after a request.

---

**Web Figure 1.**
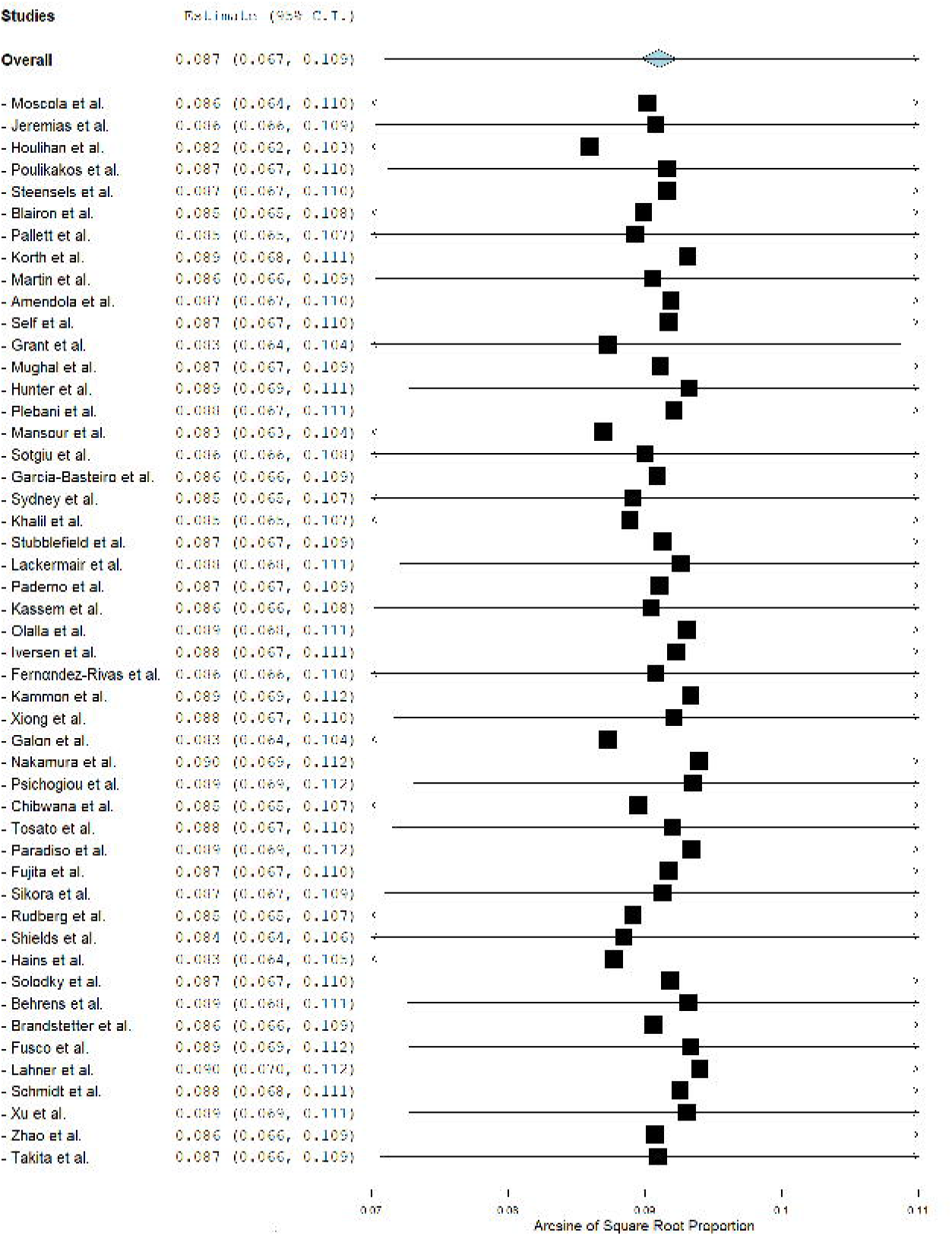
A leave-one-out sensitivity analysis of the seroprevalence of SARS-CoV-2 antibodies with corresponding 95% confidence intervals.

**Web Figure 2.**
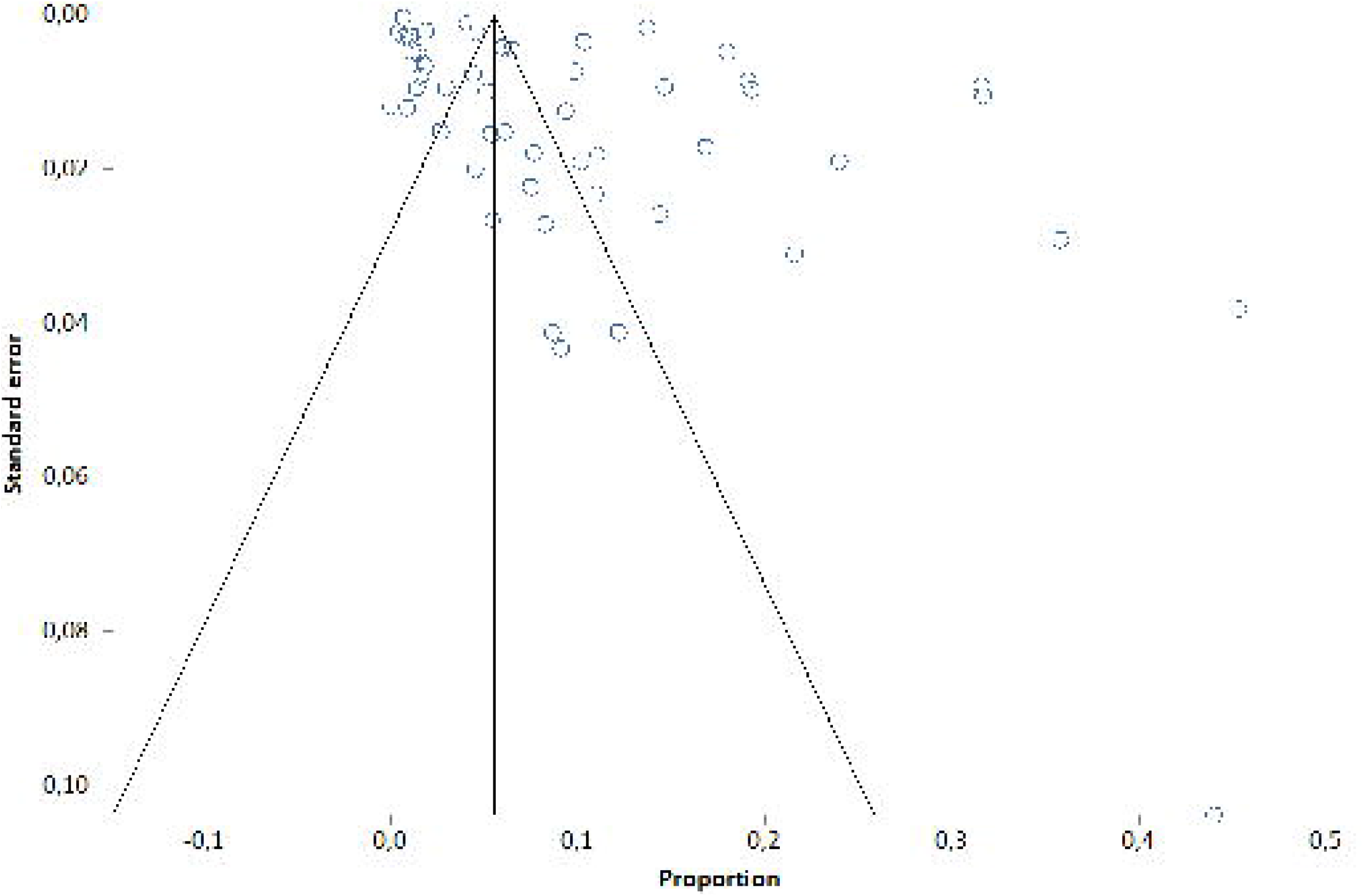
Funnel plot of the meta-analysis.

