## Supplementary material for "Seroprevalence of SARS-CoV-2 antibodies and associated factors in health care workers: a systematic review and meta-analysis": Web Tables

Web Table 1. PRISMA Checklist

| **Section/topic** | **#** | **Checklist item** | **Reported on page #** |
| --- | --- | --- | --- |
| **TITLE** | | |  |
| Title | 1 | Identify the report as a systematic review, meta-analysis, or both. | 1 |
| **ABSTRACT** | | |  |
| Structured summary | 2 | Provide a structured summary including, as applicable: background; objectives; data sources; study eligibility criteria, participants, and interventions; study appraisal and synthesis methods; results; limitations; conclusions and implications of key findings; systematic review registration number. | 1 |
| **INTRODUCTION** | | |  |
| Rationale | 3 | Describe the rationale for the review in the context of what is already known. | 3 |
| Objectives | 4 | Provide an explicit statement of questions being addressed with reference to participants, interventions, comparisons, outcomes, and study design (PICOS). | 3 |
| **METHODS** | | |  |
| Protocol and registration | 5 | Indicate if a review protocol exists, if and where it can be accessed (e.g., Web address), and, if available, provide registration information including registration number. | NA |
| Eligibility criteria | 6 | Specify study characteristics (e.g., PICOS, length of follow-up) and report characteristics (e.g., years considered, language, publication status) used as criteria for eligibility, giving rationale. | 4 |
| Information sources | 7 | Describe all information sources (e.g., databases with dates of coverage, contact with study authors to identify additional studies) in the search and date last searched. | 4 |
| Search | 8 | Present full electronic search strategy for at least one database, including any limits used, such that it could be repeated. | 4 |
| Study selection | 9 | State the process for selecting studies (i.e., screening, eligibility, included in systematic review, and, if applicable, included in the meta-analysis). | 4 |
| Data collection process | 10 | Describe method of data extraction from reports (e.g., piloted forms, independently, in duplicate) and any processes for obtaining and confirming data from investigators. | 4 |
| Data items | 11 | List and define all variables for which data were sought (e.g., PICOS, funding sources) and any assumptions and simplifications made. | 4 |
| Risk of bias in individual studies | 12 | Describe methods used for assessing risk of bias of individual studies (including specification of whether this was done at the study or outcome level), and how this information is to be used in any data synthesis. | 4-5 |
| Summary measures | 13 | State the principal summary measures (e.g., risk ratio, difference in means). | 5 |
| Synthesis of results | 14 | Describe the methods of handling data and combining results of studies, if done, including measures of consistency (e.g., I^2^) for each meta-analysis. | 5 |
| Risk of bias across studies | 15 | Specify any assessment of risk of bias that may affect the cumulative evidence (e.g., publication bias, selective reporting within studies). | 5 |
| Additional analyses | 16 | Describe methods of additional analyses (e.g., sensitivity or subgroup analyses, meta-regression), if done, indicating which were pre-specified. | 5 |
| **RESULTS** | | |  |
| Study selection | 17 | Give numbers of studies screened, assessed for eligibility, and included in the review, with reasons for exclusions at each stage, ideally with a flow diagram. | 6 |
| Study characteristics | 18 | For each study, present characteristics for which data were extracted (e.g., study size, PICOS, follow-up period) and provide the citations. | 6-7 |
| Risk of bias within studies | 19 | Present data on risk of bias of each study and, if available, any outcome level assessment (see item 12). | Web tables 3, 4, and 5 |
| Results of individual studies | 20 | For all outcomes considered (benefits or harms), present, for each study: (a) simple summary data for each intervention group (b) effect estimates and confidence intervals, ideally with a forest plot. | Figure 2 |
| Synthesis of results | 21 | Present results of each meta-analysis done, including confidence intervals and measures of consistency. | 7-8 |
| Risk of bias across studies | 22 | Present results of any assessment of risk of bias across studies (see Item 15). | 7, 8 |
| Additional analysis | 23 | Give results of additional analyses, if done (e.g., sensitivity or subgroup analyses, meta-regression [see Item 16]). | 8, Web Figure 1 |
| **DISCUSSION** | | |  |
| Summary of evidence | 24 | Summarize the main findings including the strength of evidence for each main outcome; consider their relevance to key groups (e.g., healthcare providers, users, and policy makers). | 10-13 |
| Limitations | 25 | Discuss limitations at study and outcome level (e.g., risk of bias), and at review-level (e.g., incomplete retrieval of identified research, reporting bias). | 13-14 |
| Conclusions | 26 | Provide a general interpretation of the results in the context of other evidence, and implications for future research. | 14 |
| **FUNDING** | | |  |
| Funding | 27 | Describe sources of funding for the systematic review and other support (e.g., supply of data); role of funders for the systematic review. | 14 |

Web Table 2. Validity assessment (sensitivity and specificity) for the antibodies tests used in the included studies according to the manufacturers data.

| **Reference** | **Antibodies test kit** | **Sensitivity (%)** | **Specificity (%)** |
| --- | --- | --- | --- |
| Moscola et al. 2020 [14] | EUROIMMUN^TM^  AnshLabs SARS-CoV-2 IgG ELISA  ARCHITECT, Abbott  Ortho Clinical Diagnostics VITROS Anti-SARS-CoV-2 IgG  Ortho Clinical Diagnostics VITROS Immunodiagnostic Products Anti-SARS-CoV-2 Total Reagent Pack and Calibrator Liaison (Diasorin)  Roche Elecsys Anti-SARS-CoV-2 | 90  95  100  87.5  83.3  98  100 | 100  98.3  99.6  100  100  99  99.8 |
| Jeremias et al. 2020 [15] | EUROIMMUN^TM^ | 90 | 100 |
| Houlihan et al. 2020 [16] | SARS-CoV-2 IgG ELISA | 92 | NR |
| Poulikakos et al. 2020 [17] | MAGLUMI 2019-nCoV IgM/IgG kit | 87.7-98.6 | 80.5-94.5 |
| Steensels et al. 2020 [18] | COVID-19 IgG/IgM Rapid Test Cassette; Multi-G | 71.2 | 88.3 |
| Blairon et al. 2020 [19] | Liaison (Diasorin) | 98 | 99 |
| Pallett et al. 2020 [20] | EDI Novel Coronavirus COVID-19 IgG ELISA kit | 98.4 | 99.8 |
| Korth et al. 2020 [21] | EUROIMMUN^TM^ | 90 | 100 |
| Martin et al. 2020 [22] | EUROIMMUN^TM^ | 90 | 100 |
| Amendola et al. 2020 [23] | EUROIMMUN^TM^ | 90 | 100 |
| Self et al. 2020 [24] | SARS-CoV-2 spike protein ELISA | 96 | 99 |
| Grant et al. 2020 [25] | Roche Elecsys Anti-SARS-CoV-2 | 100 | 99.8 |
| Mughal et al. 2020 [26] | STANDARD Q COVID-19 IgM/IgG Duo, SD Biosensor | 96.2 | 96.6 |
| Hunter et al. 2020 [27] | ARCHITECT, Abbott | 100 | 99.6 |
| Plebani et al. 2020 [28] | MAGLUMI 2019-nCoV IgM/IgG kit | 87.7-98.6 | 80.5-94.5 |
| Mansour et al. 2020 [29] | SARS-CoV-2 spike protein ELISA | NR | NR |
| Sotgiu et al. 2020 [30] | BioMedomics IgM-IgG Combined Antibody Rapid Test | 88.7 | 90.6 |
| Garcia-Basteiro et al. 2020 [31] | MAGPLEX, Luminex | 75 | 100 |
| Sydney et al. 2020 [32] | ARCHITECT, Abbott | 100 | 99.6 |
| Khalil et al. 2020 [33] | Abbott SARS-CoV-2 IgG ELISA | 96.9 | 99.9 |
| Stubblefield et al. 2020 [34] | SARS-CoV-2 spike protein ELISA | 96 | 99 |
| Lackermair et al. 2020 [35] | EUROIMMUN^TM^ | 90 | 100 |
| Paderno et al. 2020 [36] | Liaison (Diasorin) | 98 | 99 |
| Kassem et al. 2020 [37] | Artron Laboratories, COVID-19 IgM/IgG antibody rapid test | 93.4 | 97.7 |
| Olalla et al. 2020 [38] | COVID-19 IgG/IgM Rapid Test Cassette | 94 | 95 |
| Iversen et al. 2020 [39] | Livzon Diagnostics, COVID-19 IgM/IgG antibodies | 90.6 | 99.2 |
| Hains et al. 2020 [40] | COVID-19 Human IgM, IgG Rapid Test | 90 | 100 |
| Solodky et al. 2020 [41] | Toda Coronadiag® | 100 | 100 |
| Behrens et al. 2020 [42] | EUROIMMUN^TM^ | 90 | 100 |
| Brandstetter et al. 2020 [43] | EUROIMMUN^TM^ | 90 | 100 |
| Fusco et al. 2020 [44] | MAGLUMI 2019-nCoV IgM/IgG kit | 87.7-98.6 | 80.5-94.5 |
| Lahner et al. 2020 [45] | Commercial chemiluminesce immunoassay (CLIA), Medical Systems | 50 | 99 |
| Schmidt et al. 2020 [46] | EUROIMMUN^TM^ | 90 | 100 |
| Xu et al. 2020 [47] | Bioscience Co. magnetic chemiluminescence enzyme immunoassay (MCLIA) | NR | NR |
| Zhao et al. 2020 [48] | SARS-CoV-2 IgG ELISA | 97 | 98 |
| Fernández-Rivas et al. 2020 [49] | Liaison (Diasorin) | 98 | 99 |
| Kammon et al. 2020 [50] | One Step Novel Coronavirus (COVID-19) IgM/IgG Antibody Test | 86 | 99 |
| Xiong et al. 2020 [51] | SARS-CoV-2 IgG ELISA | NR | NR |
| Galán et al. 2020 [52] | COV19G.CE.192 | 98 | NR |
| Nakamura et al. 2020 [53] | ARCHITECT, Abbott | 100 | 99.6 |
| Psichogiou et al. 2020 [54] | GenBody COVID-19 IgM/IgG | 92 | 95 |
| Chibwana et al. 2020 [55] | Mologic SARS-CoV-2 IgG ELISA | 97 | 97 |
| Tosato et al. 2020 [56] | MAGLUMI 2019-nCoV IgM/IgG kit | 87.7-98.6 | 80.5-94.5 |
| Paradiso et al. 2020 [57] | VivaDiag^TM^ | 88 | 90 |
| Fujita et al. 2020 [58] | COVID-19 IgG ELISA kit (DRG international) | 100 | 100 |
| Sikora et al. 2020 [59] | Sugentech SGTi-flex COVID-19 IgM/IgG | 91 | 96 |
| Rudberg et al. 2020 [60] | Luminex Corp | 99 | 99 |
| Shields et al. 2020 [61] | SARS-CoV-2 spike protein ELISA | NR | NR |
| Takita et al. 2020 [62] | Kurabo Industries Ltd COVID-19 IgG | 96.7 | 95 |

NR: not reported

Web Table 3. Quality of prevalence studies.

|  | 17 | 26 | 33 | 38 | 40 | 41 | 42 | 43 | 45 | 46 | 48 | 49 | 50 | 51 | 53 | 55 | 56 | 57 | 59 | 62 |
| --- | --- | --- | --- | --- | --- | --- | --- | --- | --- | --- | --- | --- | --- | --- | --- | --- | --- | --- | --- | --- |
| 1. Was the sample frame appropriate to address the target population? | X | X |  | X | X | X | X | X | X | X |  | X |  | X | X | X | X | X | X | X |
| 2. Were study participants sampled in an appropriate way? |  |  |  |  |  |  |  |  | X | X |  |  |  |  |  |  | X |  |  |  |
| 3. Was the sample size adequate? |  |  |  |  |  |  | X | X | X | X |  |  | X |  | X |  | X | X |  |  |
| 4. Were the study subjects and the setting described in detail? |  | X |  | X | X |  |  |  | X | X |  | X |  | X | X | X | X | X | X |  |
| 5. Was the data analysis conducted with sufficient coverage of the identified sample? | X | X | X | X | X | X | X | X | X | X | X | X | X | X | X | X | X | X | X | X |
| 6. Were valid methods used for the identification of the condition? |  | X | X | X | X | X | X | X | X | X | X | X | X | X | X | X | X |  | X | X |
| 7. Was the condition measured in a standard, reliable way for all participants? | X | X | X | X | X | X | X | X | X | X | X | X | X | X | X | X | X | X | X | X |
| 8. Was there appropriate statistical analysis? | X |  |  | X | X | X | X |  |  | X | X | X |  | X | X |  | X | X | X | X |
| 9. Was the response rate adequate, and if not, was the low response rate managed appropriately? | X |  | X |  |  | X | X | X | X | X | X | X | X | X |  | X | X |  |  | X |

Web Table 4. Quality of cross-sectional studies.

|  | 14 | 15 | 18 | 19 | 21 | 23 | 24 | 25 | 26 | 28 | 29 | 30 | 31 | 32 | 34 | 35 | 36 | 37 | 39 | 44 | 47 | 52 | 54 | 58 | 60 | 61 |
| --- | --- | --- | --- | --- | --- | --- | --- | --- | --- | --- | --- | --- | --- | --- | --- | --- | --- | --- | --- | --- | --- | --- | --- | --- | --- | --- |
| 1. Were the criteria for inclusion in the sample clearly defined? | X | X | Χ | X | X | Χ | Χ | Χ | Χ |  |  |  | Χ | Χ | Χ | Χ | Χ | Χ | Χ | Χ | Χ | Χ | X | Χ | Χ | Χ |
| 2. Were the study subjects and the setting described in detail? | X | X | Χ | X | X | Χ | Χ |  | Χ |  |  | Χ | Χ | Χ | Χ | Χ | Χ | Χ | Χ | Χ | Χ | Χ | X | Χ | Χ | Χ |
| 3. Was the exposure measured in a valid and reliable way? | X | X | Χ | X | X | Χ | Χ | Χ |  | Χ | X | Χ | Χ | Χ | Χ | Χ | Χ | Χ | Χ | Χ | Χ | Χ | X | Χ | Χ | Χ |
| 4. Were objective, standard criteria used for measurement of the condition? | X | X | Χ | X | X | Χ | Χ |  | Χ |  | X | Χ | Χ | Χ | Χ | Χ | Χ | Χ | Χ | Χ | Χ | Χ | X | Χ | Χ | Χ |
| 5. Were confounding factors identified? | X |  | Χ |  |  |  |  |  |  |  |  |  | Χ |  |  |  |  |  | Χ |  |  |  |  |  |  |  |
| 6. Were strategies to deal with confounding factors stated? | X |  | Χ |  |  |  |  |  |  |  |  |  | Χ |  |  |  |  |  | Χ |  |  |  |  |  |  |  |
| 7. Were the outcomes measured in a valid and reliable way? | X | X | Χ | X | X | Χ | Χ | Χ | Χ | Χ | X | Χ | Χ | Χ | Χ | Χ | Χ | Χ | Χ | Χ | Χ | Χ | X | Χ | Χ | Χ |
| 8. Was appropriate statistical analysis used? | X | X | Χ | X |  | Χ | Χ | Χ | Χ | Χ | X | Χ | Χ |  | Χ |  | Χ | Χ | Χ |  | Χ | Χ | X | Χ | Χ | Χ |

Web Table 5. Quality of cohort studies.

|  | 16 | 20 | 22 |
| --- | --- | --- | --- |
| 1. Were the two groups similar and recruited from the same population? | Χ | Χ | Χ |
| 2. Were the exposures measured similarly to assign people to both exposed and unexposed groups? | Χ | Χ | Χ |
| 3. Was the exposure measured in a valid and reliable way? | Χ | Χ | Χ |
| 4. Were confounding factors identified? | Χ |  |  |
| 5. Were strategies to deal with confounding factors stated? |  |  |  |
| 6. Were the groups/participants free of the outcome at the start of the study (or at the moment of exposure)? | Χ | Χ |  |
| 7. Were the outcomes measured in a valid and reliable way? | Χ | Χ | Χ |
| 8. Was the follow up time reported and sufficient to be long enough for outcomes to occur? | Χ | Χ | Χ |
| 9. Was follow up complete, and if not, were the reasons to loss to follow up described and explored? | Χ | Χ |  |
| 10. Were strategies to address incomplete follow up utilized? | Χ | Χ |  |
| 11. Was appropriate statistical analysis used? | Χ | Χ | Χ |
